# Distinguishing Social Isolation, Social Support, and Contextual False Positives in Clinical Notes Using Fine-Tuned Language Models: Multisite Validation Study

**DOI:** 10.64898/2026.07.05.26357334

**Authors:** Lokesh K. Chinthala, Cindy K. Lemon, Arash Shaban-Nejad, Gregory Farage, Robert L. Davis, Hua Xu, Charisse Madlock-Brown

## Abstract

**Background:** Social isolation and social support are clinically important but inconsistently represented in structured electronic health record data. Their identification from clinical narratives is complicated by nuanced, context-dependent language and by variation in how social connectedness is documented across clinical settings, patient populations, and geographic locations. Models intended for health care use must therefore distinguish social isolation from documented support, negation, ambiguous social circumstances, and non-social clinical uses of similar terminology while remaining robust to differences in documentation practices.

**Objective:** This study aimed to develop and evaluate locally deployable, fine-tuned language models for context-aware classification of social isolation, social support, and contextually similar non-relevant clinical text across multiple health systems.

**Materials and Methods:** We conducted a retrospective study using electronic health record data from 326,847 adults aged 50 years or older who received care at 3 Tennessee health systems from 2020 to 2023. Candidate clinical text was identified using social-environment ICD-10-CM Z codes and an established social isolation lexicon. Human annotators classified 9,748 spans from 9,578 clinical notes contributed by 6,881 patients as social isolation, social support, or no social isolation. The annotated corpus included social isolation and support concepts including negations, infection-control and microbiology terminology, institutional living arrangements, and other contextually ambiguous examples. FLAN-T5-Large, BERT, RoBERTa, and Gemma-2-2B were fine-tuned for 3-class span classification. Full fine-tuning and parameter-efficient fine-tuning using low-rank adaptation approaches were evaluated for the best performing model, FLAN-T5-Large. Performance was assessed using site-based internal-external cross-validation, with macro-F1 as the primary metric.

**Results:** The annotated corpus included 2,318 social isolation spans, 638 social support spans, and 6,792 spans classified as no social isolation. Fully fine-tuned FLAN-T5-Large achieved the best performance, with a mean macro-F1 of 0.90 (SD 0.01). Class-specific F1 scores were 0.85 (SD 0.03) for social isolation, 0.87 (SD 0.04) for social support, and 0.96 (SD 0.02) for no social isolation. Gemma-2-2B achieved a macro-F1 of 0.85 (SD 0.01), compared with 0.84 (SD 0.06) for RoBERTa and 0.86 (SD 0.06) for BERT. Full fine-tuning also outperformed parameter-efficient fine-tuning in secondary pooled analyses (macro-F1 0.93 vs 0.90) in the corresponding held-out evaluation for the FLAN-T5-Large model. Remaining errors primarily involved patients who lived alone but received assistance, family presence without meaningful support, grief-related language, and non-social clinical uses of isolation terminology.

**Conclusions:** Fine-tuned language models can distinguish social isolation, social support, and contextually similar but unrelated language across heterogeneous clinical settings. FLAN-T5-Large provided the strongest and most stable performance, including for the less frequent social support class. The 3-class annotation framework, inclusion of difficult negative examples, and multisite evaluation offer a practical approach for converting nuanced social-connectedness documentation into structured data. Further work should evaluate end-to-end identification in unselected clinical notes and prospective integration into clinical and population health workflows.

## BACKGROUND AND SIGNIFICANCE

Social isolation among older adults is a significant yet underrecognized health concern, partly because relevant indicators are often buried in unstructured Electronic Health Records (EHR).[1] Defined by the relative absence of meaningful interpersonal relationships, social isolation leads to diminished social support, removing the protective buffer against negative events and reducing the benefits gleaned from positive experiences.[2] Among older adults living with serious illness, nearly one in five experience social isolation, yet fewer than 5% have this condition documented in the medical record.[3] This gap highlights the need for scalable approaches to identify social isolation directly from clinical text.

Demographic shifts, including declining family sizes, lower marriage and birth rates, and rising one-person households have intensified social isolation.[4] One-person households have grown from 13% in 1960 to 29% in 2022,[4] and the proportion of adults reporting three or fewer close friends has grown markedly, rising from approximately 27% to nearly 49% over a thirty-year period, reaching its highest level in 2021.[4]

Older adults are particularly vulnerable due to increased reliance on social support systems, making timely identification of social isolation critical for care planning and intervention. However, social isolation remains largely invisible in structured EHR data, as it is inconsistently documented and often expressed through nuanced, context-dependent language in clinical notes.

Traditional machine learning models have limited ability to capture contextual and semantic complexity. In contrast, transformer-based NLP models, including large language models (LLMs), a class of generative AI,[5] offer improved capability to identify a variety of Social Determinants of Health (SDoH) categories, interpret subtle linguistic patterns, and infer social context from within unstructured text. Recent studies have demonstrated the potential of LLMs to extract SDoH factors from clinical narratives. For example, one 2024 study used LLMs to classify six key SDoH categories (employment, housing, transportation, parental status, relationship, and social support) from the EHR text[6] with a strong performance of macro-F10.71 for any SDoH and 0.70 for adverse SDoH. Similarly, a 2025 multi-institutional study annotated 21 SDoH factors and achieved strong performance using instruction-tuned LLMs for extraction.[7] Further, researchers developed a rule-based system alongside a fine-tuned FLAN-T5-XL model to identify instances of social isolation and social support in psychiatry notes.[8] More recently, JAMIA published SDoH-GPT framework,[9] which demonstrated scalable annotation and domain adaptation. Prior work focused on broad SDoH categories with limited attention to reliable identification of social isolation and its integration into clinical workflow.

## OBJECTIVE

This study aimed to develop and evaluate a locally deployable language model fine-tuned on a diverse annotated corpus of spans extracted from clinical notes obtained from multiple specialties and care settings. The clinical notes for annotation were identified using ICD-10-CM Z-codes and established lexicons.[1] To improve model precision, we annotated false positives (e.g., negated terms, microbiology labs) and documented indicators of social support to enable meaningful comparisons between patients with and without isolation. We fine-tuned transformer-based models, including FLAN-T5-Large,[10] BERT,[11] RoBERTa,[12] and Gemma-2-2B[13] to extract social isolation concepts from unstructured clinical notes. To further improve the annotation efficiency, we incorporated an active learning framework using uncertainty sampling to prioritize the most informative examples for manual labeling during corpus development. Active learning was used to support the annotation process. However, it was not used in the model training, cross-validation, and model evaluation steps.

This work seeks to extract nuanced social context from clinical narratives to support early identification of socially isolated patients, enable population-level analyses, and inform data-driven interventions to improve outcomes and reduce disparities. One of our key design goals is ensuring the final models can be deployed locally within healthcare systems to meet privacy and regulatory constraints.

## MATERIALS AND METHODS

This observational retrospective cohort study was approved by the University of Tennessee Health Science Center (UTHSC) Institutional Review Board (IRB# 24-10073-XM). Data were obtained from the UTHSC Research Enterprise Data Warehouse (rEDW), which contains EHR data on 4.8 million patients from multiple Tennessee hospitals since 01/01/2014.[14] For this study, we used EHR data along with de-identified clinical notes from 326,847 adults aged ≥50 years between 2020-2023, across three Healthcare Organizations (HCOs): Methodist LeBonheur Healthcare (MLH), St Thomas (STH), and University of Tennessee Medical Center (UTMC), based in Memphis, Nashville, and Knoxville, TN, respectively. This cohort contributed 1,123,195 clinical notes. Analyses were conducted on the University of Tennessee’s High-performance computing cluster (ISSAC secure enclave),[15] which provides the infrastructure necessary to process Protected Health Information (PHI).

### Data

To build a high-quality training corpus, we curated a dataset of spans extracted from clinical narratives belonging to patients assigned specific SDoH ICD-10-CM Z codes,[16] including Z60.2 (Problems related to living alone), Z60.4 (Social exclusion and rejection), Z60.9 (Problems related to social environment, unspecified), Z63.2 (Inadequate family support), and Z63.3 (Absence of family member). These codes, selected with input from clinical experts, served as proxies for social isolation and guided the retrieval of notes more likely to contain relevant language. To increase representation, we supplemented the corpus with candidate spans extracted from clinical notes identified using the lexicon developed by Zhu et al.[1] This targeted sampling strategy increased the density of relevant text spans, enhancing annotation efficiency.

From 9,578 clinical notes, annotators identified and labeled 9,748 spans as ‘social isolation’ (2,318), ‘no social isolation’ (6,792), and ‘social support’ (638). Spans are sentence fragments that capture text relevant to the target concepts. Notes were collected across various departments and care settings, including the emergency department, behavioral health, and inpatient services to maximize variation in documentation style and clinical context.

Examples of core annotation spans are shown in **Supplementary Table S1**. Additional ambiguous or context-dependent spans, including institutional living, COVID-related isolation, estranged family situations, partial support patterns, and non-isolation uses of the word ‘isolated’ are provided in **Supplementary Table S2**.

Though patient and note-level characteristics are reported for cohort description, all model development and performance evaluations were conducted at the span-level.

### Annotation process

Because social isolation is context–dependent, we framed the task as span–level classification to capture precise cues and reduce noise compared to full note labeling, which often contains mixed or unrelated content. Spans extracted from clinical notes were independently annotated by two annotators using Doccano,[17] an open-source tool for text classification and sequence labelling. Notes were segmented into spans and labelled into three categories: ‘social isolation’, ‘social support’, and ‘no social isolation’. Potential false positives (e.g., pathology reports containing the term “isolated” or procedural instructions mentioning “isolation”) were explicitly annotated as ‘no social isolation’ to minimize misclassification.

Annotators followed guidelines with regular consensus meetings to ensure quality and resolve ambiguities related to span boundaries and labels. Any unresolved disagreements were reviewed by a senior domain expert. Inter-annotator agreement (IAA) was assessed using both Cohen’s kappa and percent agreement. Operational definitions included:

### social isolation

Clear indications that the patient lacks social contact or support. (e.g., No one will pick him up because he has no friends)

### social support

Explicit evidence of available emotional or practical support. (e.g., from family, friends, neighbors, or caregivers).

### no social isolation

Uses of “isolation” unrelated to social context. (e.g., microbiology isolate, infection-control isolation) or absence of any mention of social isolation or support in the text.

The annotated dataset was then used to fine-tune FLAN-T5-Large, BERT, RoBERTa, and Gemma-2-2B. Targeted annotation of both true and false positives, including negations and ambiguous cases, improved model robustness.

To improve annotation efficiency, we explored an active learning framework using uncertainty sampling. Active learning was used to prioritize candidate spans for manual review and annotation. With Monte Carlo (MC) Dropout,[18] for each unlabeled span, multiple stochastic predictions (T = 3) were generated, and model confidence was estimated from the most frequent predicted class. Spans with the lowest confidence were prioritized for manual annotation, focusing expert effort on ambiguous or informative cases. This strategy streamlined dataset expansion and improved scalability without compromising data quality.

After the annotated corpus was created, the active learning process was discontinued. The finalized annotated corpus was used for model training, validation, and testing. Therefore, all reported performance metrics were obtained using the finalized manually annotated corpus.

For patient-level descriptive summaries, patients with both social isolation and social support spans were classified as ‘social support’. This was because detecting positive social connections is often more informative for intervention and care coordination decisions. The workflow is summarized in **Figure 1**.[19]

**Figure 1.**
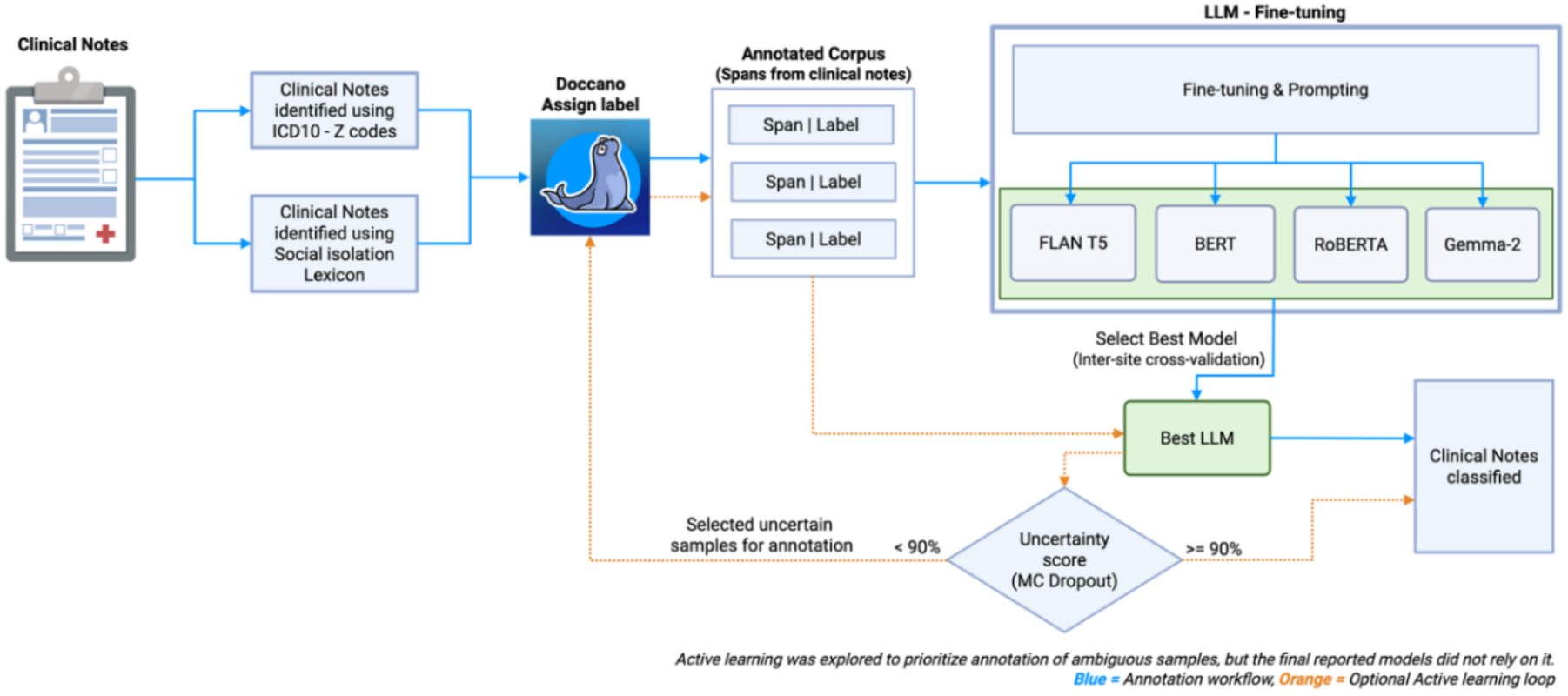
Workflow for LLM Fine-tuning with Manual Annotation and Review Loop

### Data Preprocessing

Preprocessing removed non-informative artifacts such as special characters. Text spans were extracted using annotation offsets and aligned with their corresponding labels after cleaning to ensure accurate span boundaries. No normalization was applied to preserve clinical context and linguistic variation. Duplicate records with same span and labels were dropped from the raw data before train-test split.

### Train-Test Split

All models were trained and evaluated at the span-level, consistent with the annotation schema. To assess generalizability across healthcare sites, our primary evaluation relied on site-based internal-external cross-validation (IECV), in which an entire healthcare site was held out for testing. Within each IECV fold, data from two healthcare systems were used for training and the remaining healthcare system was held out for evaluation. The training data was split into train and validation datasets. Model checkpoints were selected based on performance on the validation set and automatically reloaded at the end of training. Performance metrics were then computed on the held-out site for that fold, and results were summarized across all folds.

For the secondary analysis, the span-level annotated corpus from the three healthcare sites were combined and then divided into 80% training set, 20% test set. The 80% training set was further split into 80% train and 20% validation set for internal validation during fine-tuning. The combined multisite analyses used span-level partitioning rather than patient-level partitioning because the objective of this study was span classification rather than patient-level prediction. Therefore, the performance metrics quantify the model’s ability to classify individual annotated spans.

### Oversampling

To address class imbalance, we applied class-balanced random oversampling with replacement in the training set. Minority classes were randomly duplicated until each class contained the same number of training instances. The test set was left unchanged to avoid data leakage and ensure unbiased evaluation.

### LLM workflow

**FLAN-T5** (Fine-tuned Language Net – Text-to-Text Transfer Transformer), developed by Google Research, reframes NLP tasks as text-to-text problems[10] and was instruction-tuned for strong generalization. For this study, we used FLAN-T5-Large variant (780M parameters), which offers a strong balance of accuracy and efficiency.

**BERT** (Bidirectional Encoder Representations from Transformers) is an encoder-only model, developed by Google.[11] It introduced deep bidirectional text understanding and was pretrained using a masked language modeling (MLM) objective, predicting masked words from context, and a next sentence prediction (NSP) task to capture inter-sentence relationships.

**RoBERTa (**Robustly Optimized BERT Approach), developed by Facebook AI,[12] builds on the BERT architecture. It removes the NSP objective, replaces static masks with dynamic masking, and was trained on a larger dataset with optimized hyperparameters. These changes improved performance across standard NLP benchmarks such as GLUE[20] and SQuAD.[21]

**Gemma-2-2B** is a decoder-only lightweight model developed by Google.[13] It uses knowledge distillation to learn reasoning patterns from a larger ensemble teacher model in addition to standard training data.[22] This approach allows Gemma–2–2B to outperform larger models (20B to 30B parameters). The architecture incorporates interleaving local-global attention to reduce memory usage while retaining broader contextual understanding.[23,24] In this study, we used Gemma-2-2B (2–billion–parameter) model.

### PEFT - LoRA

Because the base models were trained on general corpora, they were fine-tuned with and without Parameter-Efficient Fine-Tuning (PEFT)[25] methods. In standard fine-tuning, the entire weight matrix is updated, which can lead to catastrophic forgetting[26] and requires large computational resources. To address this, we tested LoRA (Low-Rank Adaptation),[25] a type of PEFT method that reduces memory footprint and training overhead while preserving generalization.

For our experiments, LoRA was configured with rank r=16, scaling factor *α*=32, and dropout=0.05. The rank controlled how much task specific information could be captured, the scaling factor balanced LoRA updates with frozen weights, and dropout regularized the adapter layers to reduce overfitting. For example, instead of training 32,768 parameters for a 512×64 weight matrix, LoRA decomposes it into two smaller matrices (512×16 and 16×64), requiring only 9,216 parameters. These low-rank updates are added to the frozen weights in the self-attention layer or encoder of the transformer architecture, reducing memory usage and accelerating fine-tuning.

### Training setup

All models were fine–tuned with consistent hyperparameters: 5 epochs, with evaluation and checkpointing at the end of each epoch, a batch size of 8 and a learning rate of 5×10^-5^ were used for both training and evaluation due to GPU memory constraints, and weight decay of 0.01 was applied to reduce overfitting by penalizing large weights.

### Prompt engineering

We used structured prompts to clearly frame the classification task and reduce ambiguity, especially for FLAN–T5–Large.

Prompt engineering example –

*Classify the text into exactly one category: ‘social isolation’, ‘no social isolation’ or ‘social support’*

*Text: {text}*

*Category:*

### Site-Based Generalizability Assessment

The primary evaluation focused on model generalizability across health systems using site-based Internal-External Cross-Validation (IECV).[27] In each fold, data from two healthcare organizations were used for training and the remaining health system was held out for evaluation. This process was repeated across all three possible site combinations (MLH, STH, and UTMC), and performance metrics were summarized as the mean and standard deviation across folds.

### Combined Multisite Evaluation

Following IECV, additional analyses were conducted using the combined multisite dataset to evaluate the effect of class imbalance and compare full fine-tuning with parameter-efficient approaches when all available training data were pooled. However, these additional analyses were conducted using the overall best performing model from primary analysis – FLAN-T5-Large.

### Performance metrics

Model performance was evaluated using accuracy, precision, recall, and F1–score. Accuracy measures the proportion of correct predictions. Precision measures how many predicted positives were true positives, which is important when false positives carry high cost. Recall (sensitivity) captures how many actual positive cases were correctly identified, reducing missed detections. F1–score combines precision and recall to balance both considerations.

## RESULTS

### Participants

We curated a dataset of 9,748 labeled spans extracted from 9,578 clinical notes belonging to 6,881 patients across three health systems: MLH (n=2,661), STH (n = 2,388), and UTMC (n = 1,832), yielding a diverse study population with varying demographic distributions and documentation practices. Prevalence of social isolation differed by site: MLH (24.2%), STH (15.1%), and UTMC (52.7%). Conversely, ‘no social isolation’ was the majority class at each site, most notably at STH (81.8%). ‘Social support’ was less common, ranging from 74 patients at STH to 137 at UTMC. STH had the highest number of clinical notes – 4,285, followed by MLH with 3,081 and UTMC with 2,212 notes. Inter-annotator agreement for the combined dataset, measured using Cohen’s kappa, was 0.81, indicating substantial agreement between two annotators.

### Descriptive data

As shown in **Table 1**, demographics varied across sites. UTMC participants were on average older (mean age 72.3±11.4 years) compared to MLH (68.1±11.2) and STH (61.8±8.0). Female representation ranged from 52.1% to 54.1% across sites. Racial distributions differed substantially, with MLH having the largest proportion of African American patients (44.1%), UTMC predominantly White (91.6%), and STH was more diverse. Ethnic representation was predominantly non-Hispanic, though missingness/unknown status was highest at STH (35.8%). **Table 1**. Characteristics of the annotated population across sites

**Table 1.** Characteristics of the annotated popul ation across sites.

|  | **MLH** | **STH** | **UTMC** | **Total** |
|---|---|---|---|---|
| Study population (N) | 2,661 | 2,388 | 1,832 | 6,881 |
| social isolation, n (%) | 643 (24.2) | 361 (15.1) | 966 (52.7) | 1,970 (28.6) |
| no social isolation, n (%) | 1,882 (70.7) | 1,953 (81.8) | 729 (39.8) | 4,564 (66.3) |
| social support, n (%) | 136 (5.1) | 74 (3.1) | 137 (7.5) | 347 (5.0) |
| Demographics |  |  |  |  |
| Age, mean±SD | 68.1±11.2 | 61.8±8.0 | 72.3±11.4 | 67.0±11.1 |
| Female, n (%) | 1,387 (52.1) | 1,278 (53.5) | 992 (54.1) | 3,657 (53.1) |
| Race, n (%) |  |  |  |  |
| White | 1,426 (53.6) | 1,817 (76.1) | 1,679 (91.6) | 4,922 (71.5) |
| African American | 1,173 (44.1) | 351 (14.7) | 108 (5.9) | 1,632 (23.7) |
| Other/Unknown | 61 (2.3) | 220 (9.2) | 45 (2.5) | 326 (4.7) |
| Ethnicity, n (%) |  |  |  |  |
| Hispanic or Latino | 30 (1.1) | 74 (3.1) | 8 (0.4) | 112 (1.6) |
| Not Hispanic or Latino | 2,614 (98.2) | 1,458 (61.1) | 1,709 (93.3) | 5,781 (84.0) |
| Other/Unknown | 17 (0.6) | 856 (35.8) | 115 (6.3) | 988 (14.3) |
| Clinical Notes | 3,081 | 4,285 | 2,212 | 9,578 |

### Primary Result

The primary evaluation (**Table 2**) used site-based internal-external cross-validation to assess model generalizability across health systems. **Table 2** summarizes model performance across classes and sites. Among the four models, the fully fine-tuned FLAN-T5-Large achieved the highest and most consistent performance, with a macro-F1 of 0.90±0.01. It demonstrated strong and balanced results across classes, especially for ‘social isolation’ (F1 **=** 0.85±0.03), ‘no social isolation’ (F1 **=** 0.96±0.02), and ‘social support’ (F1 **=** 0.87±0.04). BERT obtained a macro-F1 of 0.86±0.06, performing well for ‘no social isolation’ (F1 = 0.93±0.04) but showing reduced recall and higher variability for ‘social support’ (F1 = 0.85±0.06). However, it had the highest recall of 0.90±0.10 for ‘social isolation’ compared to other models. RoBERTa achieved moderate results with a macro-F1 of 0.84±0.06, performing comparably on ‘social isolation’ and ‘no social isolation’, but with less stability across categories. Gemma-2-2B demonstrated good performance, although slightly lower than FLAN-T5-Large and BERT, with an overall macro-F1 of 0.85±0.01 and recall of 0.82±0.06 for ‘social isolation’. Overall, FLAN-T5-Large outperformed BERT, RoBERTa, and Gemma-2-2B, indicating greater generalization and robustness for social context classification in clinical notes.

**Table 2.** Site-based internal-external cross-validation performance across three health systems (Values are shown as mean±SD across folds.)

| Model | Class | Precision | Recall | F1 |
|---|---|---|---|---|
| FLAN-T5-Large | social isolation | 0.83 ± 0.07 | 0.88 ± 0.09 | **0.85 ± 0.03** |
|  | no social isolation | 0.98 ± 0.02 | 0.95 ± 0.06 | 0.96 ± 0.02 |
|  | social support | 0.88 ± 0.06 | 0.88 ± 0.07 | **0.87 ± 0.04** |
|  | macro-F1 |  |  | **0.90 ± 0.01** |
| BERT | social isolation | 0.75 ± 0.20 | **0.90 ± 0.10** | 0.80 ± 0.11 |
|  | no social isolation | 0.98 ± 0.02 | 0.89 ± 0.09 | 0.93 ± 0.04 |
|  | social support | 0.89 ± 0.02 | 0.81 ± 0.13 | 0.85 ± 0.06 |
|  | macro-F1 |  |  | 0.86 ± 0.06 |
| RoBERTa | social isolation | 0.80 ± 0.03 | 0.82 ± 0.05 | 0.81 ± 0.02 |
|  | no social isolation | 0.97 ± 0.01 | 0.96 ± 0.04 | 0.97 ± 0.02 |
|  | social support | 0.76 ± 0.16 | 0.75 ± 0.19 | 0.76 ± 0.18 |
|  | macro-F1 |  |  | 0.84 ± 0.06 |
| Gemma-2-2B | social isolation | 0.79 ± 0.07 | 0.82 ± 0.06 | 0.80 ± 0.06 |
|  | no social isolation | 0.96 ± 0.03 | 0.95 ± 0.05 | 0.96 ± 0.02 |
|  | social support | 0.80 ± 0.12 | 0.80 ± 0.06 | 0.79 ± 0.04 |
|  | macro-F1 |  |  | 0.85 ± 0.01 |

### Secondary Result

To evaluate the effect of class imbalance and parameter-efficient training, additional experiments were conducted using the combined multisite dataset. These analyses were performed using the FLAN-T5-Large model.

**Table 3** shows that fully fine-tuned FLAN-T5-Large model performed similarly with and without oversampling of minority classes, with differences primarily in precision and recall.

**Table 3.** Performance of the fully fine-tuned FLAN-T5-Large model on the combined multisite dataset with and without oversampling of minority classes.

|  | FLAN-T5-Large trained on imbalanced dataset |  |  | FLAN-T5-Large trained on balanced dataset |  |  | Support (n) |
|---|---|---|---|---|---|---|---|
|  | Precision | Recall | F1 | Precision | Recall | F1 |  |
| social isolation | **0.93** | 0.85 | 0.89 | 0.88 | **0.91** | 0.90 | 209 |
| no social isolation | 0.97 | 0.97 | 0.97 | 0.99 | 0.94 | 0.96 | 488 |
| social support | 0.85 | 0.94 | 0.89 | 0.87 | 0.97 | 0.92 | 159 |
| macro avg | 0.91 | 0.92 | 0.91 | 0.91 | 0.94 | **0.93** | 856 |

Oversampling improved social isolation recall from 0.85 to 0.91 while decreasing precision from 0.93 to 0.88, resulting in a modest improvement in macro-F1 from 0.91 to 0.93. Based on these results, the oversampled FLAN-T5-Large model was selected for subsequent comparison with the PEFT-LoRA approach. Therefore, the full fine-tuning results reported in **Table 4** correspond to the best-performing oversampled FLAN-T5-Large configuration from **Table 3**.

**Table 4.** Comparison of full fine-tuning and PEFT-LoRA fine-tuning of FLAN-T5-Large on the combined multisite dataset.

| Class | FLAN-T5-Large without PEFT-LoRA |  |  | FLAN-T5-Large with PEFT-LoRA |  |  | Support (n) |
|---|---|---|---|---|---|---|---|
|  | Precision | Recall | F1 | Precision | Recall | F1 |  |
| social isolation | 0.88 | **0.91** | **0.90** | 0.83 | 0.90 | 0.86 | 209 |
| no social isolation | 0.99 | 0.94 | 0.96 | 0.99 | 0.93 | 0.96 | 488 |
| social support | 0.87 | 0.97 | **0.92** | 0.85 | 0.89 | 0.87 | 159 |
| macro avg | 0.91 | 0.94 | **0.93** | 0.89 | 0.91 | 0.90 | 856 |

**Table 4** compares the best performing fully fine-tuned FLAN-T5-Large model with its PEFT-LoRA variant. The fully fine-tuned FLAN-T5 model achieved higher overall performance, with a macro-F1 score of 0.93 compared with 0.90 for PEFT-LoRA. For ‘social isolation’ class, fully fine-tuned FLAN-T5 achieved precision=0.88, recall=0.91, and F1= 0.90, whereas PEFT-LoRA achieved precision = 0.83, recall = 0.90, and F1= 0.86. Although PEFT-LoRA maintained reasonable recall for all three classes – ‘social isolation’ (0.90), ‘no social isolation’ (0.93), and ‘social support’ (0.89), its performance was inconsistent, particularly for social isolation (precision = 0.83, F1 = 0.86). Overall, these findings suggest that while PEFT-LoRA offers computational efficiency, full fine-tuning delivers substantially better performance for reliably classifying social isolation and related constructs.

### Error Analysis

Error analysis on a random subset of misclassified spans (Supplementary **Table S3**) revealed several patterns. First, mentions of living alone were sometimes over–interpreted as indicators of social isolation, even when support was present – “He has been living alone but states he has been self-sufficient in activities of daily living.” Second, the presence of family members occasionally led the model to predict social support despite clear evidence of limited involvement, such as “The patient’s daughter also mentioned due to past interactions she would not be able to care for the patient.” Additional errors were observed from emotional or grief–related language and from clinical uses of the term “isolated.” Although this was not a comprehensive review, these examples illustrate the contextual ambiguities that remain challenging for the model.

## DISCUSSION

The principal finding of this study is that a fine–tuned FLAN–T5–Large model with a structured prompt substantially improved detection of ‘social isolation’ and ‘social support’ within clinical notes, reducing false positives and generalizing well across diverse settings. The model generalized across three independent health systems, achieving a mean macro-F1 of 0.90 (SD 0.01) under site-based internal-external cross-validation. These results indicate that the model effectively captured subtle, context–dependent expressions of social connectedness.

Performance (macro-F1 = 0.93) remained consistent when training and evaluation were performed on the combined multisite dataset. A major contribution of this work is the improved identification of multiple social connectedness concepts by incorporating annotated examples of true and false positives, including negations and ambiguous cases. Training on both social isolation and social support enabled clearer differentiation between related constructs and improved patient stratification.

Our findings align with and extend recent efforts to extract SDoH factors, including social isolation. Patra et al. tackled the identification of social isolation and social support in psychiatric notes using both a rule-based system (RBS) and a fine-tuned FLAN-T5-XL model, reporting that the rule–based approach outperformed the LLM on macro-F1 (0.85-0.89 vs 0.65-0.82) when detecting social support and isolation categories, because it closely followed annotation guidelines, whereas the LLM’s interpretations were more liberal.[8] In contrast, our strategy fine-tuned an instruction-tuned FLAN-T5 with LoRA adapters on domain-specific data, enabling flexible language understanding while adapting to annotation rules. This resulted in performance matching or exceeding prior BERT/RoBERTa-based models reported in the literature. For example, Han et al. used a BERT classifier for multi-label SDoH classification in MIMIC-III notes and reported a best micro-F1 of 0.69.[28] An instruction-tuned LLaMA-based LLM performed at a micro-F1>0.90 on a broad SDoH classification task, underscoring the importance of fine-tuning LLMs to achieve state-of-the-art accuracy in extracting social risk factors.[7]

Our findings further complement recent work by Guevara et al., where they examined six broad SDoH categories using fine-tuned FLAN-T5 models and synthetic data augmentation.

Their best models achieved macro-F1 scores of 0.70-0.71,[6] whereas our social isolation/support-specific approach achieved substantially higher and balanced performance across classes. Additionally, unlike their focus on synthetic data and bias robustness, our workflow incorporates explicit false-positive annotation, and internal-external cross-validation across three health systems.

The SDoH-GPT by Consoli et al. focused on reducing the annotation time by using few-shot LLM prompting with various examples and concise instructions to auto-label text and then used them to train XGBoost classifier for SDoH classification task. They reported that this approach reduced the annotation time by ∼10x and cost by ∼20x, Cohen’s K was up to 0.92 and observed an AUROC > 0.90 on multiple datasets.[9] In contrast, our paper used span-level expert annotation, by explicitly including false positives, active learning via uncertainty sampling, and cross-site validation for social isolation prediction. The active learning framework was intended to improve annotation efficiency rather than model performance evaluation and served only as a mechanism for selecting informative examples for manual review.

Our future work could directly compare our instruction templates with SDoH-GPT’s sophisticated few-shot prompts and assess whether prompt engineering techniques can further streamline the annotation process and improve model generalization.

Another important point of comparison is the scope of clinical data and settings used across studies. Many earlier studies limited their corpora to specific institutions or note types. For instance, Han et al. drew from MIMIC-III social work notes focusing on ICU patients,[28] and Patra et al. used psychiatry notes from two hospitals,[8] whereas our dataset is more comprehensive, spanning a wider range of clinical note types and patient populations. This broad coverage means our model was exposed to more diverse ways that social isolation might be documented, from mental health assessments to primary care encounters, potentially making it more robust. Indeed, cross-domain generalizability is a known challenge. The 2022 n2c2 challenge on SDoH noted that models trained on one hospital’s data saw F1 scores drop from 0.89 to 0.77 when tested on a different site.[29] By curating data across varied settings (and iteratively expanding it), our work begins to bridge this gap. Active learning played a key role here: unlike most prior studies that rely on a static manually annotated corpus, we adopted an iterative annotation approach. This allowed us to prioritize “uncertain” cases for labeling, efficiently improving the model’s coverage of edge cases. Lybarger et al. previously demonstrated the value of active learning for SDoH extraction, introducing a novel framework to build an SDoH corpus with fewer annotations,[30] and our experience suggests that such strategies can improve annotation efficiency and facilitate corpus expansion. Compared to recent works, our study offers a broader and more data-efficient approach to extracting social isolation information from clinical notes.

Together, these contributions demonstrate a data-efficient and generalizable framework for extracting social isolation from clinical text, advancing the use of LLMs for clinically meaningful SDoH detection.

## INTERPRETATION AND CLINICAL IMPLICATIONS

Automated detection of social isolation from unstructured clinical notes has significant implications for clinical workflows, decision support, and population health management.

Because isolation is rarely documented in structured EHR fields, integrating fine-tuned LLMs into EHR pipelines enables extraction of actionable social context directly from clinical narratives. These extracted insights could be stored as structured variables or mapped to standardized SDoH codes (e.g., ICD-10-CM Z60.2, Z63.2), therefore enriching existing EHR datasets and improving their completeness for both clinical and research use. Embedding model-derived isolation indicators into clinical decision support dashboards, population registries, or discharge planning workflows would allow clinicians and social workers to receive automated alerts when documentation suggests a lack of social support or caregiving resources. Such real-time flagging supports timely referral to community services, behavioral health, or social work teams and aligns with the learning health system framework in which unstructured data continuously inform individualized care and quality improvement.

At a broader level, large-scale EHR integration of LLM-derived social isolation data enables systematic surveillance of social disconnection and its impact on outcomes such as readmissions, chronic disease progression, and mental health utilization. Aggregating these indicators across patient populations can identify geographic and demographic disparities, guiding targeted prevention strategies and equitable allocation of resources. Ultimately, transforming narrative clinical documentation into structured, actionable, and socially aware data bridges a critical gap between clinical observation and intervention, advancing both precision population health and equitable, person-centered care.

## LIMITATIONS

First, our dataset was drawn exclusively from Tennessee healthcare systems and focused on adults aged ≥50, which may limit generalizability to other regions or younger populations.

Linguistic patterns seen in this cohort may not reflect documentation practices elsewhere.

Second, identifying social isolation in clinical notes also remains challenging due to inconsistent terminology and the absence of a standardized lexicon, making keyword–based methods ineffective and potentially non–transferable across institutions. Although our annotations were carefully developed and adjudicated, the dataset may not capture all relevant expressions of isolation, leading to missed context–dependent cases.

Social isolation is complex and context-dependent, influenced by living situations, geographic context, and temporal changes in relationships, which complicates span–level classification. While we reviewed misclassified cases during quality checks and examined a random subset of misclassified spans, this review was not a full or systematic qualitative error analysis. As a result, the examples reported in the Results section reflect only a small portion of the misclassifications and may not capture the full range of contextual ambiguity. A more comprehensive qualitative error analysis remains an important direction for future work.

In the secondary analysis, the pooled multisite dataset was partitioned at the span level rather than the patient level given the span-level classification task. Because the analyses were performed at the span level, spans from the same patient could potentially appear in both training and test sets. However, the models operated only on span text and did not use patient-level information. Therefore, similarities in documentation across encounters may have contributed to modestly optimistic performance estimates.

Despite these limitations, our study benefits from a large, multisite dataset and from fine–tuning state–of–the–art LLMs, which have demonstrated strong performance in extracting social cues. These strengths enhance the robustness of our findings and support the effectiveness of our approach for identifying social isolation within clinical narratives.

## CONCLUSION

By leveraging FLAN-T5-Large with fine-tuning and structured prompts, this study substantially improved the accuracy of identifying “social isolation” and “social support” within unstructured clinical notes while reducing false positives. Comparative analyses showed that FLAN-T5-Large outperformed BERT, RoBERTa, and Gemma-2-2B in the current classification task, highlighting the advantages of instruction-tuning strategies.

To enhance computational efficiency, parameter-efficient fine-tuning (PEFT) using LoRA were also evaluated. Although the LoRA-based FLAN-T5-Large model achieved lower F1-score compared to full fine-tuning, it required significantly fewer computational resources, demonstrating a viable trade-off for resource-constrained settings. These findings indicate that both full and parameter-efficient adaptations of FLAN-T5-Large can generalize effectively across care settings and produce contextually relevant predictions that reliably capture social constructs.

Beyond methodological gains, this work demonstrates how large language models can transform free-text EHR data into actionable, structured insights that enhance detection of social risk, support targeted interventions, and inform population-level analyses. Future work will focus on integrating these models directly into EHR workflows and expanding detection to other social determinants of health to further advance equitable, data-driven care.

## Author Contributions

LC conceptualized the study, performed data curation, formal analysis, investigation, methodology development, project administration, resource management, software development, validation, and drafted and edited the manuscript. CL contributed to validation and manuscript editing. GF, RD, and HX contributed to manuscript editing. AS-N contributed to drafting and editing the manuscript. CM-B provided supervision, contributed to validation, and drafted and edited the manuscript. The authors used generative AI tools (e.g., ChatGPT, Gemini) to assist with language editing, grammar refinement and code optimization. All substantive content, analyses, and conclusions were developed by the authors.

## Data availability

The datasets generated and/or analyzed during the current study are not publicly available due to privacy issues. All codes for model fine-tuning are available on GitHub (https://github.com/lokeshchinthala/transformer-based-social-isolation-detection-nlp).

## REFERENCES

1 Zhu VJ, Lenert LA, Bunnell BE, et al. Automatically identifying social isolation from clinical narratives for patients with prostate Cancer. BMC Med Inform Decis Mak. 2019;19:43. doi: 10.1186/s12911-019-0795-y

2 Gable SL, Bedrov A. Social isolation and social support in good times and bad times. Curr Opin Psychol. 2022;44:89–93. doi: 10.1016/j.copsyc.2021.08.027

3 Ritchie CS, Kotwal AA. Loneliness and Social Isolation in Palliative Care: A Call to Action. J Palliat Med. 2023;26:1032–4. doi: 10.1089/jpm.2023.0425

4 Bureau UC. Census Bureau Releases New Estimates on America&#8217;s Families and Living Arrangements. Census.gov. https://www.census.gov/newsroom/press-releases/2022/americas-families-and-living-arrangements.html (accessed 31 July 2025)

5 Shaban-Nejad A, Michalowski M, Bianco S. Creative and generative artificial intelligence for personalized medicine and healthcare: Hype, reality, or hyperreality? Exp Biol Med. 2023;248:2497–9. doi: 10.1177/15353702241226801

6 Guevara M, Chen S, Thomas S, et al. Large language models to identify social determinants of health in electronic health records. Npj Digit Med. 2024;7:6. doi: 10.1038/s41746-023-00970-0

7 Keloth VK, Selek S, Chen Q, et al. Social determinants of health extraction from clinical notes across institutions using large language models. Npj Digit Med. 2025;8:287. doi: 10.1038/s41746-025-01645-8

8 Patra BG, Lepow LA, Kasi Reddy Jagadeesh Kumar P, et al. Extracting social support and social isolation information from clinical psychiatry notes: comparing a rule-based natural language processing system and a large language model. J Am Med Inform Assoc. 2025;32:218–26. doi: 10.1093/jamia/ocae260

9 Consoli B, Wang H, Wu X, et al. SDoH-GPT: using large language models to extract social determinants of health. J Am Med Inform Assoc. 2025;ocaf094. doi: 10.1093/jamia/ocaf094

10 Chung HW, Hou L, Longpre S, et al. Scaling Instruction-Finetuned Language Models. 2022.

11 Devlin J, Chang M-W, Lee K, et al. BERT: Pre-training of Deep Bidirectional Transformers for Language Understanding. 2019.

12 Liu Y, Ott M, Goyal N, et al. RoBERTa: A Robustly Optimized BERT Pretraining Approach. 2019.

13 Team G, Riviere M, Pathak S, et al. Gemma 2: Improving Open Language Models at a Practical Size. 2024.

14 About CBMI. UTHSC. https://uthsc.edu/cbmi/about/index.php (accessed 8 January 2026)

15 UTK O of IT. System Overview on ISAAC Secure Enclave | Office of Innovative Technologies. https://oit.utk.edu/hpsc/isaac-secure/system-overview-secure-research-enclave/ (accessed 6 May 2024)

16 IMPROVING THE COLLECTION OF Social Determinants of Health (SDOH) Data with ICD-10-CM Z Codes. https://www.cms.gov/files/document/cms-2023-omh-z-code-resource.pdf (accessed 7 August 2025)

17 Nakayama H, Kubo T, Kamura J, et al. doccano: Text Annotation Tool for Human. 2018.

18 Gal Y, Ghahramani Z. Dropout as a Bayesian Approximation: Representing Model Uncertainty in Deep Learning. 2016.

19 Created in BioRender. Chinthala, L. (2026) https://BioRender.com/nlm877j.

20 Wang A, Singh A, Michael J, et al. GLUE: A Multi-Task Benchmark and Analysis Platform for Natural Language Understanding. 2019.

21 Guven ZA, Unalir MO. Natural language based analysis of SQuAD: An analytical approach for BERT. Expert Syst Appl. 2022;195:116592. doi: 10.1016/j.eswa.2022.116592

22 Hinton G, Vinyals O, Dean J. Distilling the Knowledge in a Neural Network. 2015.

23 Beltagy I, Peters ME, Cohan A. Longformer: The Long-Document Transformer. 2020.

24 Luong M-T, Pham H, Manning CD. Effective Approaches to Attention-based Neural Machine Translation. 2015.

25 Lialin V, Deshpande V, Yao X, et al. Scaling Down to Scale Up: A Guide to Parameter-Efficient Fine-Tuning. 2024.

26 Ven GM van de, Soures N, Kudithipudi D. Continual Learning and Catastrophic Forgetting. 2024.

27 Takada T, Nijman S, Denaxas S, et al. Internal-external cross-validation helped to evaluate the generalizability of prediction models in large clustered datasets. J Clin Epidemiol. 2021;137:83–91. doi: 10.1016/j.jclinepi.2021.03.025

28 Han S, Zhang RF, Shi L, et al. Classifying social determinants of health from unstructured electronic health records using deep learning-based natural language processing. J Biomed Inform. 2022;127:103984. doi: 10.1016/j.jbi.2021.103984

29 Romanowski B, Ben Abacha A, Fan Y. Extracting social determinants of health from clinical note text with classification and sequence-to-sequence approaches. J Am Med Inform Assoc. 2023;30:1448–55. doi: 10.1093/jamia/ocad071

30 Lybarger K, Ostendorf M, Yetisgen M. Annotating social determinants of health using active learning, and characterizing determinants using neural event extraction. J Biomed Inform. 2021;113:103631. doi: 10.1016/j.jbi.2020.103631

